# Association between self-reported STI symptoms and stillbirth in sub-Saharan Africa using nationally representative household surveys

**DOI:** 10.1101/2023.08.23.23294477

**Authors:** Skylar Pulver, Casey N. Pinto, Djibril M. Ba, Kathryn A. Risher

**Affiliations:** Penn State College Medicine, Dept Public Health Sciences, Hershey, PA

## Abstract

Stillbirths are a known outcome of untreated sexually transmitted infections (STIs). Sub-Saharan Africa (SSA) shares a disproportionate burden of both stillbirths (2015: 29/1000 births vs 3/1000 births in high income countries globally) and STIs. Nationally representative survey data can inform stillbirth prevention measures in multiple countries in the SSA region, with greater generalizability than clinic-based studies. We assessed the association between any self-reported STI symptoms (STI diagnosis, abnormal genital discharge, genital sore or ulcer) in the last 12 months and stillbirth in the prior five years, among women aged 15-49 years participating in nationally representative Demographic and Health Surveys (DHS) in 19 countries in SSA between 2015-2021 and reporting a pregnancy in the five years prior to interview. We used multivariable logistic regression models adjusting for maternal age, wealth index, education level of mother, cigarette smoking behaviors, health care utilization and country of residence. Among 160,995 SSA women who reported being pregnant at least once between 2011-2021, 18.0% (25,596) reported any STI symptom in the prior year and 1.9% (3,205) reported a stillbirth in the past five years. Women who self-reported STI symptoms in the past year had 1.30 (95% CI 1.16-1.47) times the odds of stillbirth in the prior five years compared to women who did not report STI symptoms. Women 35-49 years old had 1.17 (95% CI 1.04-1.33) times the odds of stillbirth compared to those 15-24 years old. The association found between self-reported STI symptoms and stillbirth highlights the need for increased screening for STI symptoms and subsequent treatment among pregnant women in SSA.

## Introduction

The rate of stillbirth globally has been steadily decreasing over the last two decades, with the most recent UNICEF data showing a stillbirth rate of 13 per 1,000 births globally. Although this rate is decreasing, this still means that there were around 1.9 million stillbirths globally in 2021 alone (1). Stillbirth rates in sub-Saharan Africa (SSA) are higher than those in high-income countries globally with 29 stillbirths per 1,000 births, compared to 3 per 1,000 births (2). High rates of stillbirth in SSA continue to be a healthcare burden, with a recent study conducted in Zambia and Tanzania suggesting that with careful measurement, rates may be higher than current national estimates (3). An association between sexually transmitted infections (STIs) and adverse pregnancy outcomes such as spontaneous abortion, stillbirth, prematurity, and low birth weight (LBW) has been previously documented (4). Although STIs are a major global public health concern, with 1 million STIs acquired daily, countries in SSA are disproportionately affected by these infections (5). Forty percent of the 60 million newly diagnosed STIs globally are diagnosed in SSA, these diagnosed STIs can be either symptomatic or asymptomatic (6). One barrier to treatment and diagnosis is the asymptomatic nature of the majority of STIs (5). Prevention of adverse pregnancy outcomes includes testing for HIV, syphilis, and hepatitis B (HBsAg) preferably during the first trimester of pregnancy, per the World Health Organization (WHO). These recommendations do not include recommendations on clinical diagnostic testing for herpes, gonorrhea, chlamydia, or trichomoniasis (7). However, in high-resource settings, the CDC provides recommendations for women at risk to also be screened for gonorrhea and chlamydia due to potential adverse health outcomes (8). It is essential to continue to screen pregnant women at risk of STIs in SSA, even if asymptomatic, due to the high rate of STIs in this region and globally.

With the burden of disease for both STIs and stillbirth taking a toll on populations in the SSA region, the relationship between these two factors must continue to be investigated. Self-reported STIs (SR-STIs) provide one avenue to capture and potentially treat those who have not been tested in resource-limited settings. Recent research has shown the importance of laboratory diagnosis for STIs, as the majority of STIs are asymptomatic, yet in resource-limited settings this transition has yet to have been implemented (9). Therefore, analyses using only clinically diagnosed STI infections could be influenced by different types of bias than SR-STI symptoms due to lack of care seeking/ behaviors, as well as access to care.

The nationally representative Demographic and Health Surveys (DHS) are completed approximately every 5 years in low- and middle-income countries, and ask questions related to a myriad of lifestyle/health topics including maternal health and SR-STIs (10). Previous research using DHS data in SSA highlights SR-STI symptoms in this region being positively associated with sexual autonomy and negatively associated with healthcare-seeking behaviors (11, 12). This study will contribute to the literature that shows the utility of DHS data in learning more about the association between the burden of STI and stillbirths in this region.

Overall, there is a lack of research on the role of SR-STI symptoms in stillbirth in SSA. Previous studies in Kenya have determined there to be an association between diagnosed STIs such as syphilis, but not any other diagnosed STIs (13). We propose to close that gap by assessing the relationship between SR-STI symptoms on the outcome of stillbirth using DHS data. Investigating this association will help public health professionals to better understand the relationship between STIs and stillbirth, and help identify strategies for stillbirth prevention.

## Materials and Methods

### Data source and study population

The DHS are nationally representative household surveys completed approximately every 5 years in low- and middle-income countries (LMICs) with questions focusing on a range of topics including sociodemographic characteristics, health behaviors, reproductive health, and nutrition (10). SSA countries that conducted a DHS between 2015-2021 were included in this study. The 19 countries included in this study were Angola, Benin, Burundi, Cameroon, Ethiopia, Gambia, Guinea, Liberia, Madagascar, Malawi, Mali, Mauritania, Nigeria, Rwanda, Sierra Leone, South Africa, Uganda, Zambia, and Zimbabwe. We investigated the association between SR-STI symptoms in the prior year and stillbirth over the last five years before surveys were conducted. For each of these countries, data were analyzed from the most recent Standard DHS completed in each country. The study population consisted of women 15-49 years old who reported being pregnant at least once in the 5 years prior to the DHS interview. Data used in this study were downloaded from the DHS website (http://www.dhsprogram.com) after project approval from DHS.

### Dependent Variable

The dependent variable was stillbirth in the five years prior to when a participant was interviewed. The WHO defines stillbirth as a pregnancy loss after 28 weeks or 7 months, therefore in this study we defined stillbirth as a pregnancy loss at 7 months or later (14). Specific variables to measure stillbirth in the DHS are limited, therefore, data from the following questions were combined to define the stillbirth outcome. Women were asked 1.“Have you ever had a pregnancy that miscarried, was aborted, or ended in stillbirth?” (Yes, No); 2.“When did the last such pregnancy end?”; 3.“How many months pregnant were you when the last such pregnancy ended?”. These questions were combined to look specifically at those who answered all questions to meet criteria for stillbirth. In order to only look at recent stillbirths, only pregnancies in the five years prior to the surveys were used, with no data included from pregnancies before 2010.

### Independent variable

The main independent variable was self-reported STI symptoms in the last 12 months, which was created as a composite variable from a set of self-reported STI symptom questions. The following questions were used to create the composite independent variable for this study based on previously published research; 1. “During the last 12 months have you had a disease which you got through sexual contact?” (Yes, No); 2. “Sometimes women experience a bad-smelling abnormal genital discharge. During the last 12 months, have you had a bad-smelling abnormal genital discharge?” (Yes, No); 3. “Sometimes women have a genital sore or ulcer. During the last 12 months, have you had a genital sore or ulcer?” (Yes, No) (12). An indicator variable was created to include any “Yes” to the previous three questions to indicate self-reported STI symptoms in the respondent.

### Covariates

Previous studies reported that the following sociodemographic and socioeconomic factors might affect a woman’s likelihood of stillbirth and STIs in LMICs, thus we included maternal age, wealth index, education level of mother, cigarette smoking behaviors, health care utilization and country of residence as covariates in this study (15, 16). All covariates are self-reported. The wealth index is calculated by the DHS based on self-reported information collected from respondents. Wealth index were grouped by DHS into categories of ‘Poorest,’ ‘Poorer,’ ‘Middle,’ ‘Rich,’ and ‘Richest’. Households are placed in these categories using principal component analysis of households’ assets such as the types of consumer goods they own (e.g., household ownership of a television and car), sources of drinking water and sanitation services available, and type of flooring in their home (17).

### Statistical Analyses

We present descriptive tabulations of absolute counts and survey-weighted proportions for our variables of interest. Simple and multivariable logistic regression models were performed accounting for the complex survey design. DHS survey weights were adjusted to weigh each survey equally in the analysis. The covariates of maternal age, wealth index, education level of mother, cigarette smoking behaviors, health care utilization and country of residence were controlled for while the association between STI symptoms and stillbirth was investigated. The variables selected for inclusion in our multivariable regression models were selected based on a conceptual framework. The multivariable logistic regression results are presented as adjusted odds ratios (AOR) with 95% confidence intervals (CIs). In sensitivity analyses, we ran multivariable logistic regression models adjusting for the same variables within each country dataset individually, to present country-level AORs for the association between STI symptoms and stillbirth, with a random effects meta-analysis of the country-level AORs using the inverse variance method for pooling. Additionally, we repeated our same multivariable logistic regression model with all countries adjusting for the same variables using two definitions of our population of interest and outcome of interest 1) stillbirth in the prior five years among those who were pregnant in the prior five years, and 2) stillbirth in the prior year among those who were pregnant in the prior year. With these two outcomes, we present AORs for the previously described SR-STI composite measure and each of its component questions individually. We further run these models sub-setting to only those who were HIV-negative at the time of survey, among the subset of women for whom HIV status was available. All analyses were conducted using R statistical software version 4.2.2 (R Foundation for Statistical Computing, Vianna, Austria) at a two-tailed alpha level of 0.05.

### Ethical Considerations

The Penn State University IRB deemed this study to be non-human subjects’ research. Data used in this study were downloaded from the DHS website (http://www.dhsprogram.com) after project approval from DHS. More details on ethical considerations can be found in the DHS methodology, “Protecting the Privacy of DHS Survey Respondents” (18).

## Results

### Socio-demographic characteristics of participants

A total of 160,995 women with a survey-weighted mean age of 29.2 years old (SD = 7.3 years old), who reported being pregnant at least once in SSA between the years of 2011-2021 were included in this study. At the time the surveys were conducted, 45.0% (71,620) of women in this study were between 25-34 years old, and 29.7% (48,057) were 15-24 years old (Table 1). In our sample there was a slight over-representation of the lowest wealth quintile (21.2%, 37,289) and a slight under-representation of the highest wealth quintile (18.1%, 27,260). About a third (35.1%, 56,969) of the population had no education and 4.1% (6,560) of the population had higher than secondary education. Cigarette smoking was not prevalent in this population with 97.1% (157,808) of women reporting they had never smoked cigarettes. Healthcare services were utilized by 63.8% (101,371) of the population in the year prior to survey.

**Table 1.**
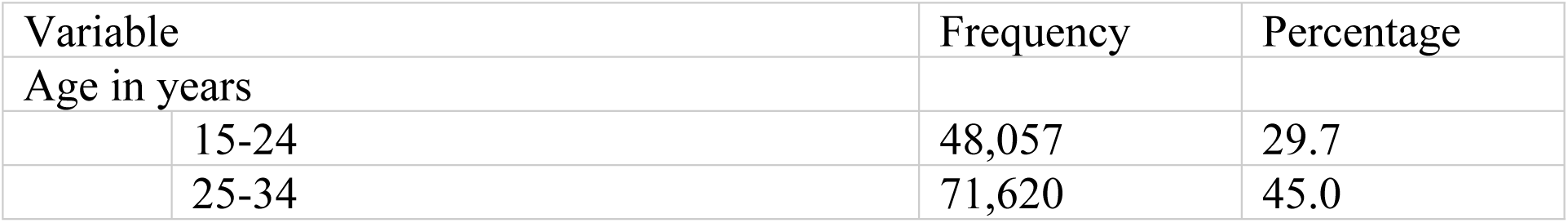
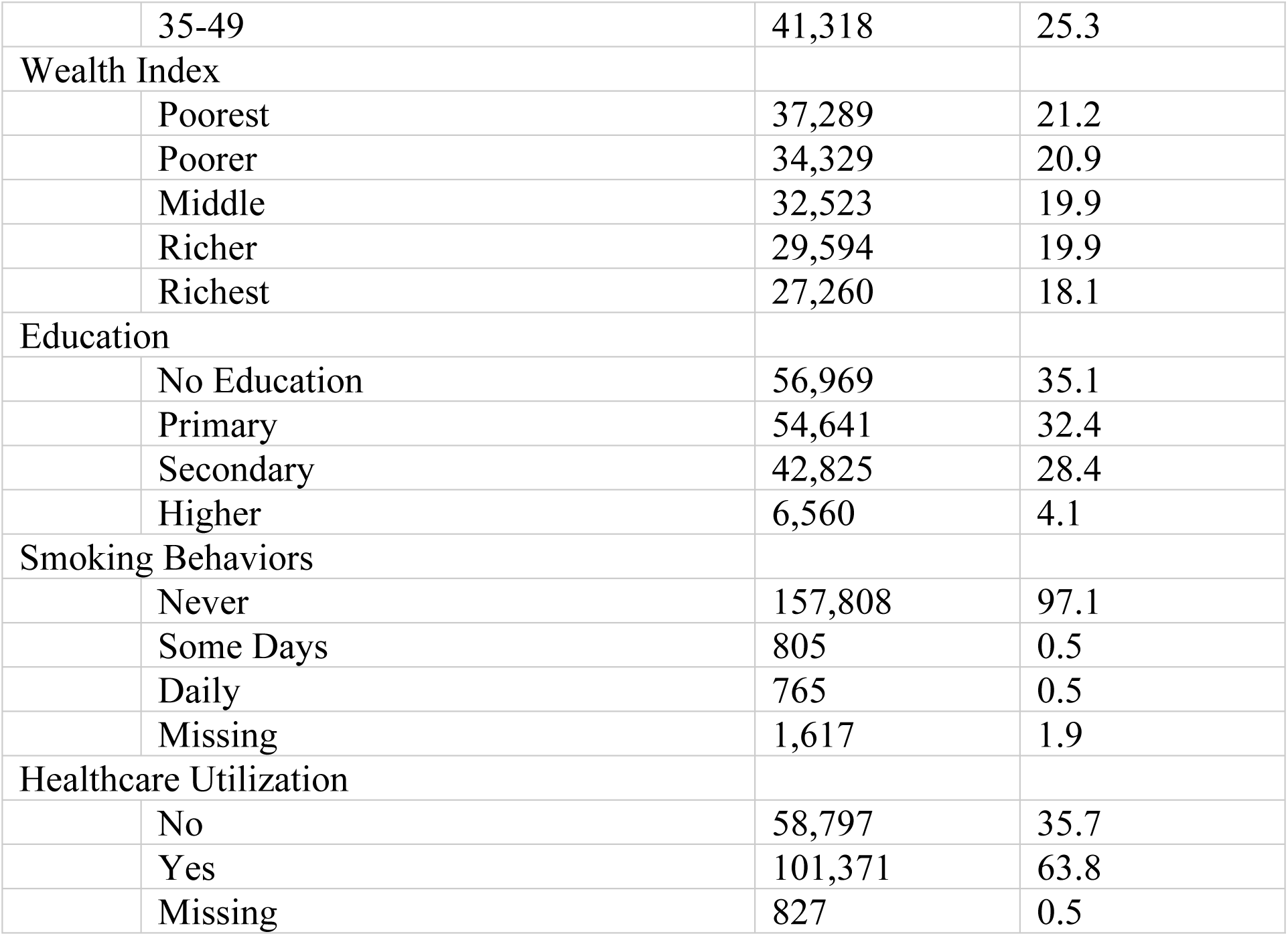
Socio-demographic characteristics of participants (N = 160,995)

| Variable | Frequency | Percentage |
| --- | --- | --- |
| Age in years |  |  |
| 15-24 | 48,057 | 29.7 |
| 25-34 | 71,620 | 45.0 |

|  |  |  |  |
| --- | --- | --- | --- |
|  | 35-49 | 41,318 | 25.3 |
| Wealth Index |  |  |  |
|  | Poorest | 37,289 | 21.2 |
|  | Poorer | 34,329 | 20.9 |
|  | Middle | 32,523 | 19.9 |
|  | Richer | 29,594 | 19.9 |
|  | Richest | 27,260 | 18.1 |
| Education |  |  |  |
|  | No Education | 56,969 | 35.1 |
|  | Primary | 54,641 | 32.4 |
|  | Secondary | 42,825 | 28.4 |
|  | Higher | 6,560 | 4.1 |
| Smoking Behaviors |  |  |  |
|  | Never | 157,808 | 97.1 |
|  | Some Days | 805 | 0.5 |
|  | Daily | 765 | 0.5 |
|  | Missing | 1,617 | 1.9 |
| Healthcare Utilization |  |  |  |
|  | No | 58,797 | 35.7 |
|  | Yes | 101,371 | 63.8 |
|  | Missing | 827 | 0.5 |

### Prevalence of Individual Self-Reported STI Symptoms (SR-STI) in sub-Saharan Africa

A disease acquired through sexual contact in the last 12 months was reported in 7.3% (10,168) of women included in the study (Table 2), while 7.8% (11,379) of women responded that they had a genital sore/ulcer in the past 12 months. Additionally, 13.9% (19,156) of women reported abnormal genital discharge in the past 12 months. Any STI symptoms (STI diagnosis, genital sore/ulcer, abnormal genital discharge) were reported in 18.0% (25,596) of the women in this study. When looking at SR-STI symptoms by country and region in SSA that were included in this study, Guinea (36.3%) and Liberia (49.4%) had the highest percentage of SR-STI symptoms in Western SSA, Cameroon (18.8%) in Central SSA, Malawi (14.7%) and Uganda (25.3%) in Eastern SSA, and South Africa (12.7%) in Southern SSA (Figure 1a.).

**Figure 1.**
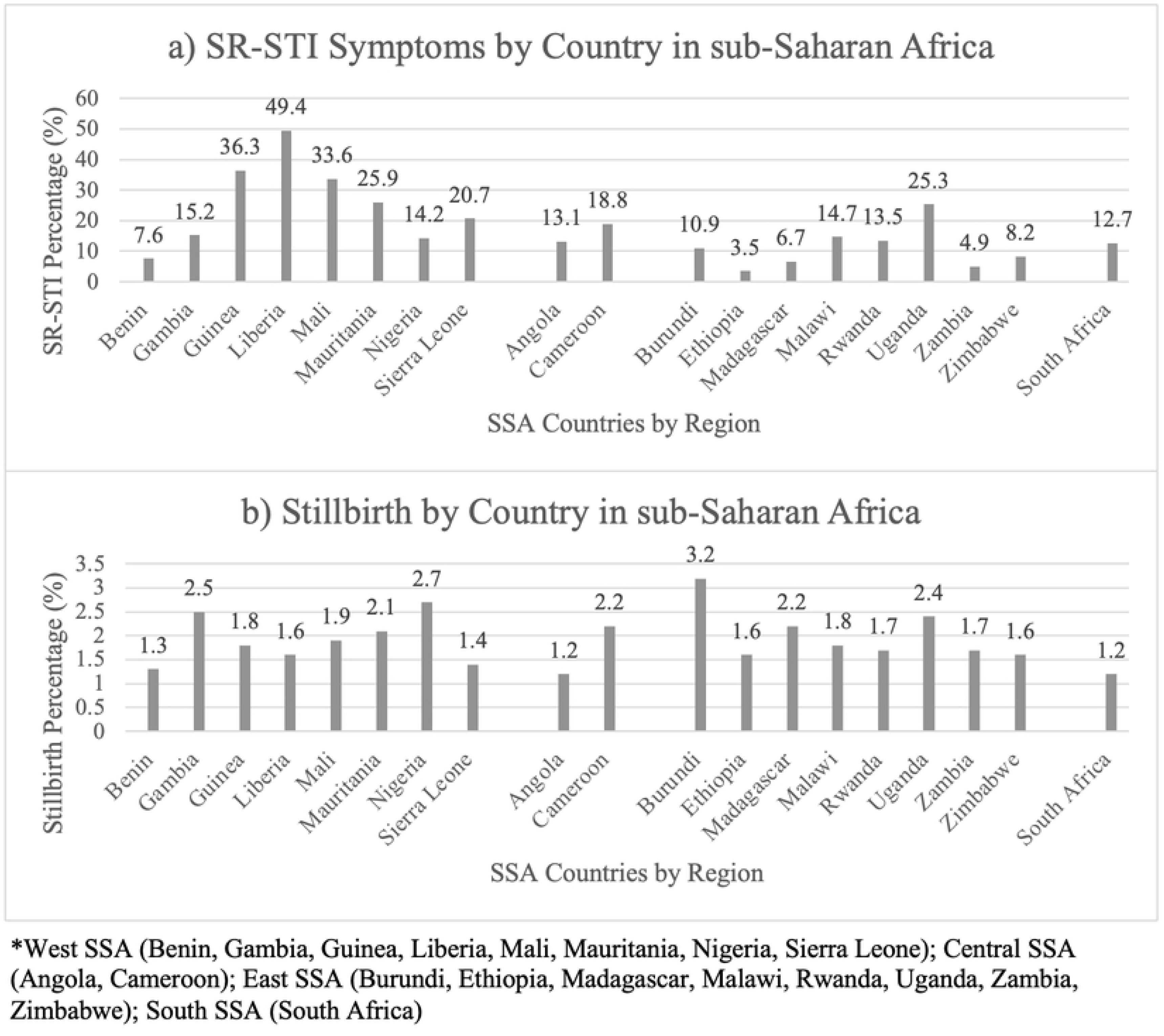
Percent of a) SR-STI Symptoms in past year and b) Stillbirth in prior 5 years by Country in sub-Saharan Africa.

**Table 2.**
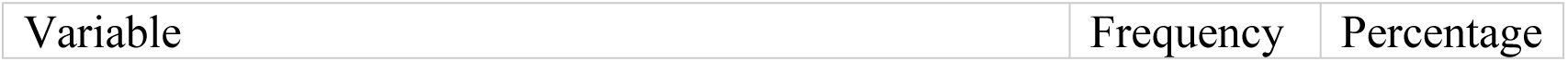
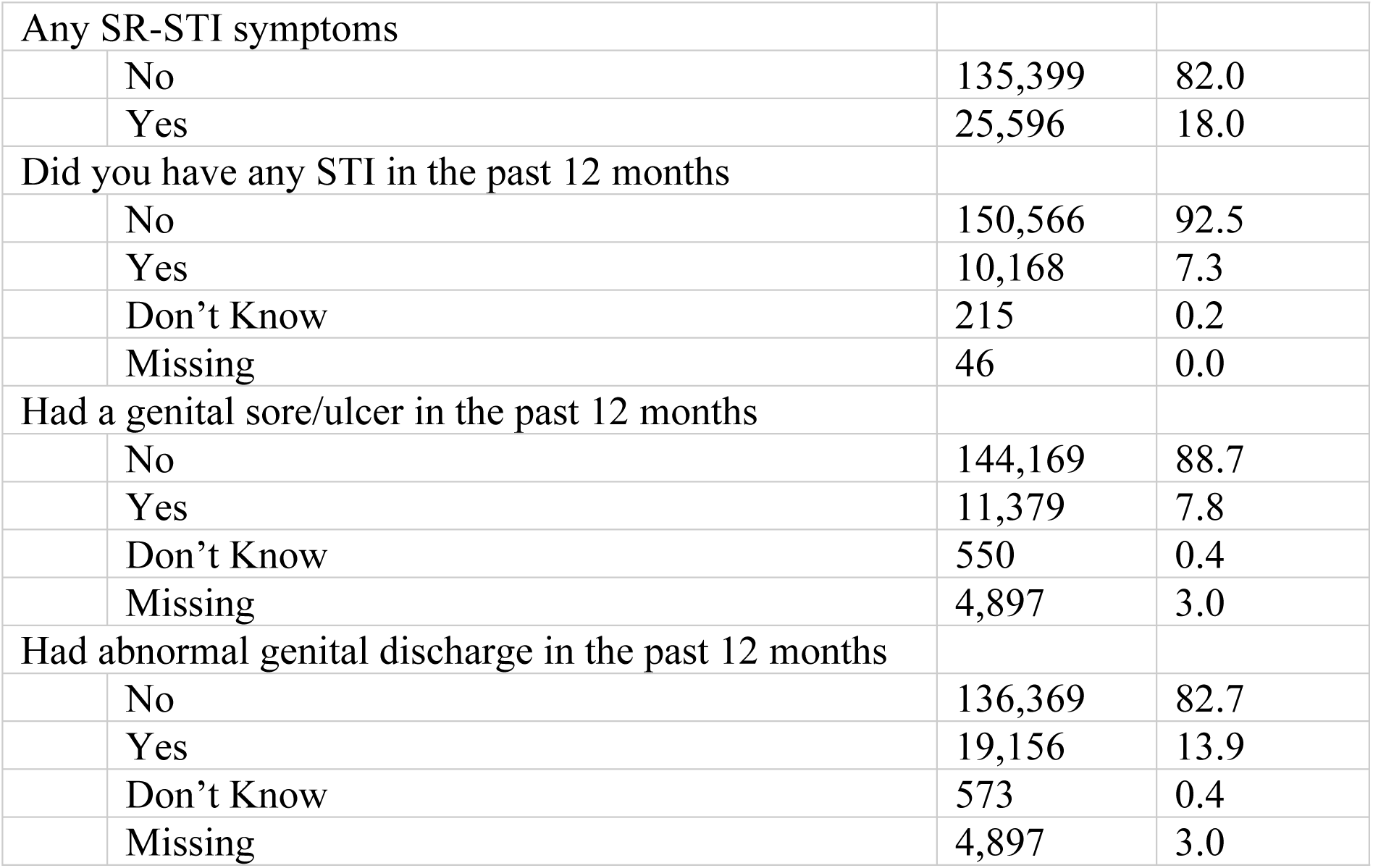
Prevalence of individual self-reported STI symptoms (N = 160,995)

| Variable | Frequency | Percentage |
| --- | --- | --- |

|  |  |  |  |
| --- | --- | --- | --- |
| Any SR-STI symptoms |  |  |  |
|  | No | 135,399 | 82.0 |
|  | Yes | 25,596 | 18.0 |
| Did you have any STI in the past 12 months |  |  |  |
|  | No | 150,566 | 92.5 |
|  | Yes | 10,168 | 7.3 |
|  | Don't Know | 215 | 0.2 |
|  | Missing | 46 | 0.0 |
| Had a genital sore/ulcer in the past 12 months |  |  |  |
|  | No | 144,169 | 88.7 |
|  | Yes | 11,379 | 7.8 |
|  | Don't Know | 550 | 0.4 |
|  | Missing | 4,897 | 3.0 |
| Had abnormal genital discharge in the past 12 months |  |  |  |
|  | No | 136,369 | 82.7 |
|  | Yes | 19,156 | 13.9 |
|  | Don't Know | 573 | 0.4 |
|  | Missing | 4,897 | 3.0 |

### Prevalence of Stillbirth in sub-Saharan Africa

Among the 19 countries in sub-Saharan Africa that were included in this study, 1.9% (3,205) of women reported the loss of a pregnancy at seven months or later in the five years prior to survey, between 2011-2021. When looking at stillbirth percentages by country and region in SSA that were included in this study, Gambia (2.5%) and Nigeria (2.7%) had the highest stillbirth percentage for Western SSA, Cameroon (2.2%) for Central SSA, Burundi (3.2%) and Uganda (2.4%) for Eastern SSA, and South Africa (1.2%) for Southern SSA (Figure 1b.).

### Factors Associated with Stillbirth in sub-Saharan Africa

In the unadjusted model, women that self-reported STI symptoms in the past year had 1.26 (crude odds ratio (COR) = 1.26, 95% CI 1.13-1.42) times the odds of having a stillbirth in the prior five years compared to women who did not report STI symptoms (Table 3). When compared to 15-24 year-old women, women aged 35-49 had 1.22 (COR = 1.22, 95% CI 1.08-1.37) times the odds of stillbirth, whereas 25-34 year-old women did not see a measurable difference in odds (COR = 0.96, 95% CI 0.86-1.07). Women in the richest quintile of the wealth index had a 15% (COR = 0.85, 95% CI 0.73-0.98) lower odds of experiencing stillbirth than those in the poorest quintile, while those in the poorer, middle, and richer quintiles did not have a measurable difference. Those with secondary education had 0.79 (COR = 0.79, 95% CI 0.70-0.90) times the odds of having a stillbirth when compared to those who had no education. There was no statistically significant difference between those with primary (COR = 0.99, 95% CI 0.90-1.11) or higher than secondary education (COR = 1.03, 95% CI 0.79-1.34) when compared to those with no education. No significant association was found between those who smoked cigarettes some days (COR = 0.99, 95% CI 0.41-2.34) and those who smoked cigarettes daily (COR = 1.06, 95% CI 0.54-2.06) compared to those who did not smoke cigarettes. The odds of having a stillbirth among women who reported utilizing healthcare services in the prior year were 1.16 (COR = 1.16, 95% CI 1.05-1.29) times the odds among women who did not.

**Table 3.** Logistic regression analysis of factors associated with stillbirth in prior five years.

| Variable | Ever had a stillbirth | COR (95% CI) | AOR (95% CI)<sup>#</sup> |
| --- | --- | --- | --- |
|  | Yes (survey prop) |  |  |
| SR-STI |  |  |  |
| No | 2,568 (1.8) | 1 | 1 |
| Yes | 637 (2.3) | 1.26 (1.13, 1.42)* | 1.30 (1.16, 1.47)* |
| Age in years |  |  |  |
| 15-24 | 944 (1.9) | 1 | 1 |
| 25-34 | 1,313 (1.8) | 0.96 (0.86, 1.07) | 0.93 (0.83, 1.04) |
| 35-49 | 948 (2.2) | 1.22 (1.08, 1.37)* | 1.17 (1.04, 1.33)* |
| Wealth Index |  |  |  |
| Poorest | 769 (1.9) | 1 | 1 |
| Poorer | 697 (2) | 1.05 (0.92, 1.19) | 1.06 (0.93, 1.21) |
| Middle | 669 (2) | 1.03 (0.91, 1.17) | 1.05 (0.92, 1.19) |
| Richer | 611 (1.9) | 0.99 (0.86, 1.14) | 1.02 (0.88, 1.19) |
| Richest | 459 (1.6) | 0.85 (0.73, 0.98)* | 0.85 (0.71, 1.00) |
| Education |  |  |  |
| No Education | 1,236 (2) | 1 | 1 |
| Primary | 1,084 (2) | 0.99 (0.90, 1.11) | 0.99 (0.88, 1.12) |
| Secondary | 770 (1.6) | 0.79 (0.70, 0.90)* | 0.88 (0.76, 1.03) |
| Higher | 115 (2.1) | 1.03 (0.79, 1.34) | 1.19 (0.88, 1.62) |
| Smoking Behaviors |  |  |  |
| Never | 3,161 (1.9) | 1 | 1 |
| Some Days | 11 (1.9) | 0.99 (0.41, 2.34) | 1.07 (0.45, 2.54) |
| Daily | 13 (2) | 1.06 (0.54, 2.06) | 1.14 (0.58, 2.24) |
| Healthcare Utilization in prior year |  |  |  |
| No | 1,065 (1.7) | 1 | 1 |
| Yes | 2,126 (2) | 1.16 (1.05, 1.29)* | 1.12 (1.01, 1.25)* |

\*p-value $\leq 0.05$
#Full model was adjusted for country of residence

After adjusting for pre-specified confounders, women who self-reported STI symptoms in the past year had 1.30 (adjusted odds ratio (AOR) = 1.30, 95% CI 1.16-1.47) times the odds of stillbirth in the prior five years compared to women who did not report STI symptoms (Table 3). Women 35-49 years old had 1.17 (AOR = 1.17, 95% CI 1.04-1.33) times the odds of stillbirth compared to those 15-24 years old. In the adjusted model no significant difference was found between women in the poorer (AOR = 1.06, 95% CI 0.93-1.21), middle (AOR = 1.05, 95% CI 0.92-1.19), richer (AOR = 1.02, 95% CI 0.88-1.19), and richest (AOR = 0.85, 95% CI 0.71-1.00) wealth index quintile when compared to those in the poorest wealth index quintile. Women with primary (AOR = 0.99, 95% CI 0.88-1.12), secondary (AOR = 0.88, 95% CI 0.76-1.03), and higher than secondary education (AOR = 1.19, 95% CI 0.88-1.62) did not have a statistically significant difference in odds of stillbirth when compared to those with no education. Where never smoking cigarettes was the baseline, no statistically significant association was found between smoking cigarettes some days (AOR = 1.07, 95% CI 0.45-2.54) and daily (AOR = 1.14, 95% CI 0.58-2.24) with chances of stillbirth. The odds of having a stillbirth among those reporting that they had utilized healthcare were 1.12 (AOR = 1.12, 95% CI 1.01-1.25) times the odds of those who did not.

A meta-analysis of individual country-adjusted regression models was conducted (Figure 2). Individual country estimates of the association (AOR) between self-reported STI in the prior year and stillbirth in the prior five years ranged from 0.44 (South Africa) to 1.89 (Zimbabwe). However, all the CIs overlapped each other, and the overall pooled adjusted odds ratio was 1.31 (95% CI: 1.17-1.46). Statistically significant results were also found in Guinea (AOR = 1.65, 95% CI 1.03-2.63), Madagascar (AOR = 1.86, 95% CI 1.12-3.10), and Nigeria (AOR = 1.44, 95% CI 1.13-1.84) for the association between SR-STI and stillbirth.

**Figure 2.**
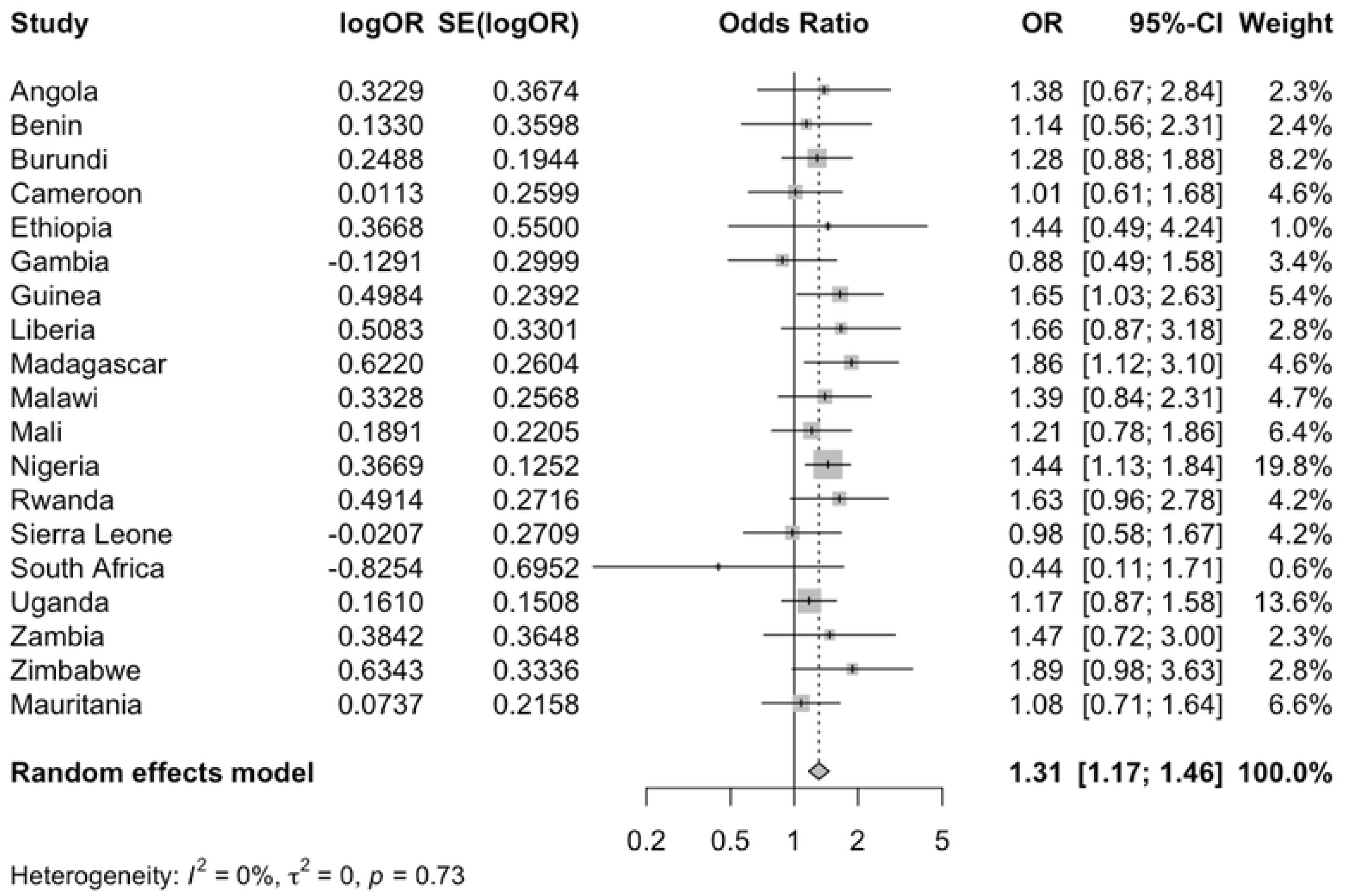
Forest Plot of Country-specific adjusted odds ratios of the association between SR-STI in prior year and stillbirth in prior five year.

Our additional sensitivity analyses (S1 Table), comparing the association of each SR-STI component question with stillbirth in the prior five years and in the prior year, among all respondents and only those HIV-negative at survey, found largely consistent results across each of our outcome definitions, SR-STI question components, and within the subset of HIV negative women.

## Discussion

This study found that women who self-reported an STI in the past 12 months were 30% more likely to have a stillbirth in the prior five years than those who did not. While the overall proportion reporting a stillbirth or SR-STI symptoms in the population was low, in terms of absolute numbers, within the study population, over 10,000 women self-reported STI symptoms in the past 12 months, and 3,205 women reported a stillbirth in the past 5 years. When adjusted to the population level, assessing for SR-STI could make a large impact on the stillbirth rate in SSA. The adjusted model in this study demonstrated that factors such as SR-STI symptoms, maternal age, and healthcare utilization are significantly associated with stillbirth in SSA. This is consistent with literature which shows there has been an association found between a high burden of STIs, such as in Liberia, and an increased risk of maternal and neonatal outcomes which include stillbirth, but also include miscarriage (12). If left untreated, 10-15% of women with STIs such as chlamydia and gonorrhea will develop pelvic inflammatory disease (PID), which can increase the risk of infertility and ectopic pregnancies (19, 20). As STIs continue to increase globally, investigations into improving SR-STI reporting and subsequent treatment can have an impact on maternal and child health in SSA where STIs pose a significant health burden (6).

There is a notable between-country variability in both SR-STI symptom prevalence and stillbirth prevalence. Liberia had the highest proportion of SR-STI (49.4%) and one of the lowest proportions of stillbirth (1.6%). Burundi had the highest stillbirth percentage of the countries included in this study at (3.2%) and a SR-STI percentage of 10.9%, much lower than Liberia. However, the country-specific AORs in these countries for the relationship between SR-STI and stillbirth both showed a positive association (AOR = 1.28 in Burundi and 1.66 in Liberia). This disparity in overall prevalence could potentially be explained by better self-awareness of STI status and symptoms among women in Liberia, and good access to care lowering stillbirth rates. Conversely women in Burundi may not have access to care overall as the political instability of the country over the last several decades has resulted in an extremely fragmented and damaged healthcare system that may result in heightened risk of stillbirths (21). Further, with 47% of women in Burundi reporting no schooling or primary schooling, a gap in health literacy regarding symptoms of STIs could explain why such low STI numbers are seen in Burundi (21). While the study population was defined differently and resulted in a different overall prevalence of stillbirth, an East African study in 2021, similarly found that Burundi had the highest burden of stillbirth among countries assessed (22).

As maternal age increases, so do the risks for pregnancy. Women in this study considered to be of advanced maternal age, between the ages of 35-49 years at study interview, had an increased risk of experiencing a stillbirth when compared to women 15-24 years old. A recent meta-analysis focusing on women of advanced maternal age showed an increased risk of ectopic pregnancy, gestational diabetes, hypertension and pre-eclampsia, cesarean delivery, maternal mortality, spontaneous miscarriage, chromosomal abnormalities, preterm delivery, and stillbirth (23). The increased burden of these factors places an additional strain on LMIC health care services. Without adequate access to healthcare services, the combination of several of these factors can lead to an increase in both stillbirth and maternal mortality.

The measure of healthcare utilization in this study provides insight into whether or not any services were accessed by the women surveyed, but not what services were accessed or available. A significant association was found among women in SSA between reporting utilizing healthcare services and experiencing a stillbirth. It should be noted that healthcare utilization was asked in the survey for the prior year, while stillbirth was for the prior five years. Women without prior complications may not seek out healthcare and may be less likely to utilize healthcare services than a woman who has experienced a stillbirth. In SSA, 31% of pregnant women deliver their children in a homebirth setting (24). In the event of complications, homebirths in LMICs have less access to lifesaving resources and skilled birth attendants. A study completed in Tanzania found that women who delivered outside a health facility were 1.85 times more likely to experience neonatal deaths when compared to those who delivered in a healthcare facility (25). Therefore, out of the women practicing homebirths, those who experience a stillbirth or miscarriage may be more likely to seek out a healthcare facility in the future, than those who did not have a miscarriage or stillbirth.

The use of DHS data for this study allows the results to be more representative of the population, while also limits the study by the questions which are asked in the questionnaire. With the use of cross-sectional data for this analysis no direct causation can be found between the variables in question. Self-reported STI symptoms were asked of the women in this study regarding the previous 12 months prior to survey completion, while the pregnancy data used to determine stillbirth was within the five years prior to the survey completion date.

The investigation into variables associated with stillbirth in this study was hampered by the limited way in which DHS asks about stillbirth in their survey. The measure of stillbirth had to be calculated during analysis as the only variable in DHS asking about stillbirth asks about miscarriage, abortion, and stillbirth all in one question. The consolidation of these three distinct pregnancy outcomes into one question limits the ability for researchers to assess the impact of these three outcomes independently and determine with accuracy the impact on global populations (26).

Recall bias is another limitation in this study, implicit in the design of the DHS which relies on participants to recall specific information about their medical history. For this analysis the variable of SR-STI symptoms required recall of 12 months. When asked about the termination of past pregnancies, women were asked to recall in which month the pregnancy was terminated. The data used in this analysis was limited to the five years prior to this survey, and participants response of the month in which their past pregnancies terminated was used to create the stillbirth variable. Although there are limitations to this study, it utilized nationally representative data to find significant associations between SR-STI symptoms and stillbirth in SSA, as well as capture populations that may not have access to diagnostic testing and are therefore often excluded from research on STIs.

## Conclusion

This study found a significant association between SR-STI symptoms and stillbirth in SSA, which could be used to target interventions in this population. In order to help reduce the global burden of stillbirths, and increase overall maternal health, we recommend an increase in discussions about STIs during pregnancy, a greater availability of testing, and an increased access to treatment. The nationally representative data used in this analysis strengthens the results of this study, and allows the results to inform broader populations than previous studies which were completed on an individual country basis. Therefore, this study informs public health professionals on areas to target with the goal of reducing the percentage of stillbirths in sub-Saharan Africa, which could impact global stillbirth prevalence.

## Data Availability

In this manuscript we conduct secondary data analysis of publicly accessible data available upon request from the Demographic and Health Survey (DHS) website (https://dhsprogram.com/data/available-datasets.cfm). Unfortunately we cannot share the data because registration through DHS is required for access to data.

## Supporting information

**Appendix A**: Supplementary table

## References

1. UNICEF. Stillbirths and stillbirth rates [Internet]. 2023 [cited 2023 May 24]. Available from: https://data.unicef.org/topic/child-survival/stillbirths/#:~:text=Around%201.9%20million%20stillbirths%20%E2%80%93%20babies,stillbirths%20per%201%2C000%20total%20births

2. Lavender DT, Chimwaza A, Daley R, Wood J [Internet]. The University of Manchester; 2022 [cited 2023 May 24]. Available from: https://sites.manchester.ac.uk/stillbirth-prevention-africa/

3. Bedwell C, Blaikie K, Actis Danna V, Sutton C, Laisser R, Tembo Kasengele C, et al. Understanding the complexities of unexplained stillbirth in sub-saharan Africa: A mixed-methods study. BJOG: An International Journal of Obstetrics & Gynaecology. 2021 Jun;128(7):1206–14. doi:10.1111/1471-0528.16629

4. Mullick S, Watson-Jones D, Beksinska M, Mabey D. Sexually transmitted infections in pregnancy: Prevalence, impact on pregnancy outcomes, and approach to treatment in developing countries. Sexually Transmitted Infections. 2005 Aug;81(4):294–302. doi:10.1136/sti.2002.004077

5. WHO. Sexually transmitted infections (STIs) [Internet]. World Health Organization; 2022 [cited 2023 May 24]. Available from: https://www.who.int/news-room/fact-sheets/detail/sexually-transmitted-infections-(stis)

6. Jarolimova J, Platt LR, Curtis MR, Philpotts LL, Bekker L-G, Morroni C, et al. Curable sexually transmitted infections among women with HIV in sub-Saharan africa. AIDS. 2022 Apr 1;36(5):697–709. doi:10.1097/qad.0000000000003163

7. WHO. Dual HIV/syphilis rapid diagnostic tests can be used as the first test in antenatal care [Internet]. World Health Organization; 2019 [cited 2023 May 24]. Available from: https://www.who.int/publications/i/item/WHO-CDS-HIV-19.38

8. CDC. Pregnancy and HIV, viral hepatitis, STD, & TB Prevention [Internet]. Centers for Disease Control and Prevention; 2022 [cited 2023 May 24]. Available from: https://www.cdc.gov/nchhstp/pregnancy/default.htm#:~:text=Prenatal%20screening%20for%20some%20infections,women%20at%20risk%20for%20infection

9. Ngobese B, Abbai NS. Sexually transmitted infections in pregnant women from sub-Saharan Africa. Southern African Journal of Infectious Diseases. 2021 Dec 9;36(1). doi:10.4102/sajid.v36i1.312

10. USAID. The DHS program [Internet]. 2023 [cited 2023 May 24]. Available from: https://dhsprogram.com/Methodology/Survey-Types/DHS.cfm

11. Adu C, Mohammed A, Budu E, Frimpong JB, Tetteh JK, Ahinkorah BO, et al. Sexual autonomy and self-reported sexually transmitted infections among women in sexual unions. Archives of Public Health. 2022 Jan 26;80(1). doi:10.1186/s13690-022-00796-4

12. Birhane BM, Simegn A, Bayih WA, Chanie ES, Demissie B, Yalew ZM, et al. Self-reported syndromes of sexually transmitted infections and its associated factors among reproductive (15– 49 years) age women in Ethiopia. Heliyon. 2021 Jul 9;7(7). doi:10.1016/j.heliyon.2021.e07524

13. Warr AJ, Pintye J, Kinuthia J, Drake AL, Unger JA, McClelland RS, et al. Sexually transmitted infections during pregnancy and subsequent risk of stillbirth and infant mortality in Kenya: a prospective study. Sexually Transmitted Infections. 2019 Feb;95(1):60–6. doi:10.1136/sextrans-2018-053597

14. WHO. Stillbirth [Internet]. World Health Organization; 2023 [cited 2023 May 24]. Available from: https://www.who.int/health-topics/stillbirth#tab=tab_1

15. Di Mario S, Say L, Lincetto O. Risk factors for stillbirth in developing countries: a systematic review of the literature. Sexually Transmitted Diseases. 2007 Jul;34(7). doi:10.1097/01.olq.0000258130.07476.e3

16. Christou A, Dibley MJ, Raynes-Greenow C. Beyond counting stillbirths to understanding their determinants in low- and middle-income countries: A systematic assessment of stillbirth data availability in household surveys. Tropical Medicine & International Health. 2016 Dec 19;22(3):294–311. doi:10.1111/tmi.12828

17. DHS. Ethiopia demographic and Health Survey 2016 [FR328] [Internet]. 2017 [cited 2023 May 24]. Available from: https://dhsprogram.com/pubs/pdf/FR328/FR328.pdf

18. DHS. The DHS program [Internet]. 2022 [cited 2023 May 24]. Available from: https://dhsprogram.com/methodology/Protecting-the-Privacy-of-DHS-Survey-Respondents.cfm

19. CDC. STD facts - pelvic inflammatory disease [Internet]. Centers for Disease Control and Prevention; 2022 [cited 2023 May 24]. Available from: https://www.cdc.gov/std/pid/stdfact-pid.htm

20. CDC. STDs & Infertility [Internet]. Centers for Disease Control and Prevention; 2023 [cited 2023 May 24]. Available from: https://www.cdc.gov/std/infertility/default.htm

21. Akombi BJ, Ghimire PR, Agho KE, Renzaho AM. Stillbirth in the African Great Lakes region: A pooled analysis of Demographic and Health Surveys. PLOS ONE. 2018 Apr 29; doi:10.1371/journal.pone.0202603

22. Tesema GA, Tessema ZT, Tamirat KS, Teshale AB. Prevalence of stillbirth and its associated factors in East Africa: generalized linear mixed modeling. BMC Pregnancy and Childbirth. 2021 Jun 2;21. doi:10.1186/s12884-021-03883-6

23. Glick I, Kadish E, Rottenstreich M. Management of Pregnancy in Women of Advanced Maternal Age: Improving Outcomes for Mother and Baby. International Journal of Women’s Health. 2021 Aug;13:751–9. doi:10.2147/ijwh.s283216

24. Hernández-Vásquez A, Chacón-Torrico H, Bendezu-Quispe G. Prevalence of home birth among 880,345 women in 67 low- and middle-income countries: A meta-analysis of Demographic and Health Surveys. SSM - Population Health. 2021 Nov 3;16:100955. doi:10.1016/j.ssmph.2021.100955

25. Ajaari J, Masanja H, Weiner R, Abokyi SA, Owusu-Agyei S. Impact of Place of Delivery on Neonatal Mortality in Rural Tanzania. International Journal of MCH and AIDS. 2012; doi:10.21106/ijma.10

26. Christou A, Dibley MJ, Raynes-Greenow C. Beyond counting stillbirths to understanding their determinants in low- and middle-income countries: A systematic assessment of stillbirth data availability in household surveys. Tropical Medicine &amp; International Health. 2017 Jan 22;22(3). doi:10.1111/tmi.12828

